# Polygenic scores for disease risk are not associated with clinical outcomes in Parkinson’s disease

**DOI:** 10.1101/2025.01.31.25321395

**Authors:** Manuela MX Tan, Hirotaka Iwaki, Sara Bandres-Ciga, Yuri Sosero, Maryam Shoai, Kathrin Brockmann, Nigel M Williams, Roy N Alcalay, Jodi Maple-Grødem, Guido Alves, Ole-Bjørn Tysnes, Peggy Auinger, Shirley Eberly, Peter Heutink, David K. Simon, Karl Kieburtz, John Hardy, Caroline H Williams-Gray, Donald G Grosset, Jean-Christophe Corvol, Ziv Gan-Or, Mathias Toft, Lasse Pihlstrøm

## Abstract

Polygenic risk scores (PRS) in Parkinson’s disease (PD) are associated with disease risk. Recently, pathway-specific PRS have been created to take advantage of annotations inking variants to biological pathways or cell types. Here, we investigated 8 biological pathways or regions of open chromatin using pathway-specific PRS: alpha-synuclein pathway, adaptive immunity, innate immunity, lysosomal pathway1, endocytic membrane-trafficking pathway, mitochondrial pathway, microglial open chromatin single nucleotide polymorphisms (SNPs), and monocyte open chromatin SNPs. We analysed 7,402 PD patients across 18 ‘in-person’ PD cohorts, and 6,717 patients from the online Fox Insight study. We did not find any significant associations between the 8 pathway-specific PRSs and 8 clinical outcomes in PD. Though this may be due to a lack of statistical power and limited sample size, it may also suggest that the genetic architecture of sporadic PD risk is different from the genetics of PD progression and clinical outcomes.

## Introduction

Parkinson’s disease (PD) is a neurodegenerative condition characterised by motor impairments and a range of non-motor features, including cognitive impairment, dementia, depression and autonomic symptoms. Genome-wide association studies (GWASs) have identified common variants that contribute to PD risk^1,2^.

Polygenic risk scores (PRSs) are a useful method of combining information from GWAS risk variants and can be used to compare individuals with high and low genetic risk. A PRS is calculated as a weighted sum of risk alleles an individual carries, with variants weighted by their effect size from a GWAS^3^. These can include just genome-wide significant variants (sometimes referred to as the Genetic Risk Score; GRS) or include also other other variants below a more liberal significance threshold.

In PD, the PRS has been shown to discriminate between PD cases and controls, with an odds ratio of 3.74 for individuals in the lowest compared to the highest quartile^1^. In addition, the PD GRS has been associated with some clinical outcomes, including younger age at onset^4^, time to Hoehn and Yahr stage 3 or greater (H&Y3)^5^, and cognitive decline and motor decline^6^. However some of these previous studies have been small and results have not been consistently replicated other than for age at onset. Recent large-scale GWASs of PD progression have consistently found no association between the PD GRS and clinical outcomes or progression^7–10^.

These studies investigated a general PD-PRS, capturing the cumulative effect of all common risk variants. In contrast, the PRS algorithm may also take advantage of annotations linking variants to biological pathways or cell types in order to generate *stratified* or *pathway-specific* PRS. Such pathway-specific PD-PRSs have been shown to be associated with disease risk in a number of previous studies^11–15^. We hypothesized that the striking clinical heterogeneity across individual PD patients reflect differences in the underlying molecular pathogenesis, associated with specific genetic vulnerabilities that may be profiled using pathway-specific PD-PRSs. This would be in line with evidence from patients with coding mutations in monogenic PD genes, where patients carrying variants in *GBA1*, involved in the lysosomal pathway, have more rapid progression^16,17^, whereas mitochondrial and mitophagy genes *PRKN* and *PINK1* are associated with slower progression and less cognitive impairment^18–23^, though more frequent dystonia^19,22^. PD patients carrying mutations in *LRRK2* have slower progression of motor symptoms^24^ and better cognitive performance than non-carriers^25^. Importantly, the same genetic mutations associated with increased PD risk can be associated with slower or faster progression. However, there may also be distinct genetic factors, which are not associated with disease risk, that influence the rate of progression and clinical _features7,9,10,26._

Our goal was to investigate the association between clinical outcomes and different pathway-specific polygenic risk scores (PRS) in Parkinson’s disease (PD). Disentangling these associations will provide important novel insights into the mechanisms shaping the phenotype of PD and facilitate patient stratification into biologically relevant subtypes for clinical trials and future precision medicine.

In order to limit multiple testing, we took a hypothesis driven approach. We investigated 8 biological pathways or regions of open chromatin which have previously been associated with PD risk (Figure 1): alpha-synuclein pathway^11^, adaptive immunity^11,12^, innate immunity^11,12^, lysosomal pathway^11,13^, endocytic membrane-trafficking pathway^14^, mitochondrial pathway^15^, microglial open chromatin SNPs^27^, and monocyte open chromatin SNPs^27^.

**Figure 1.**
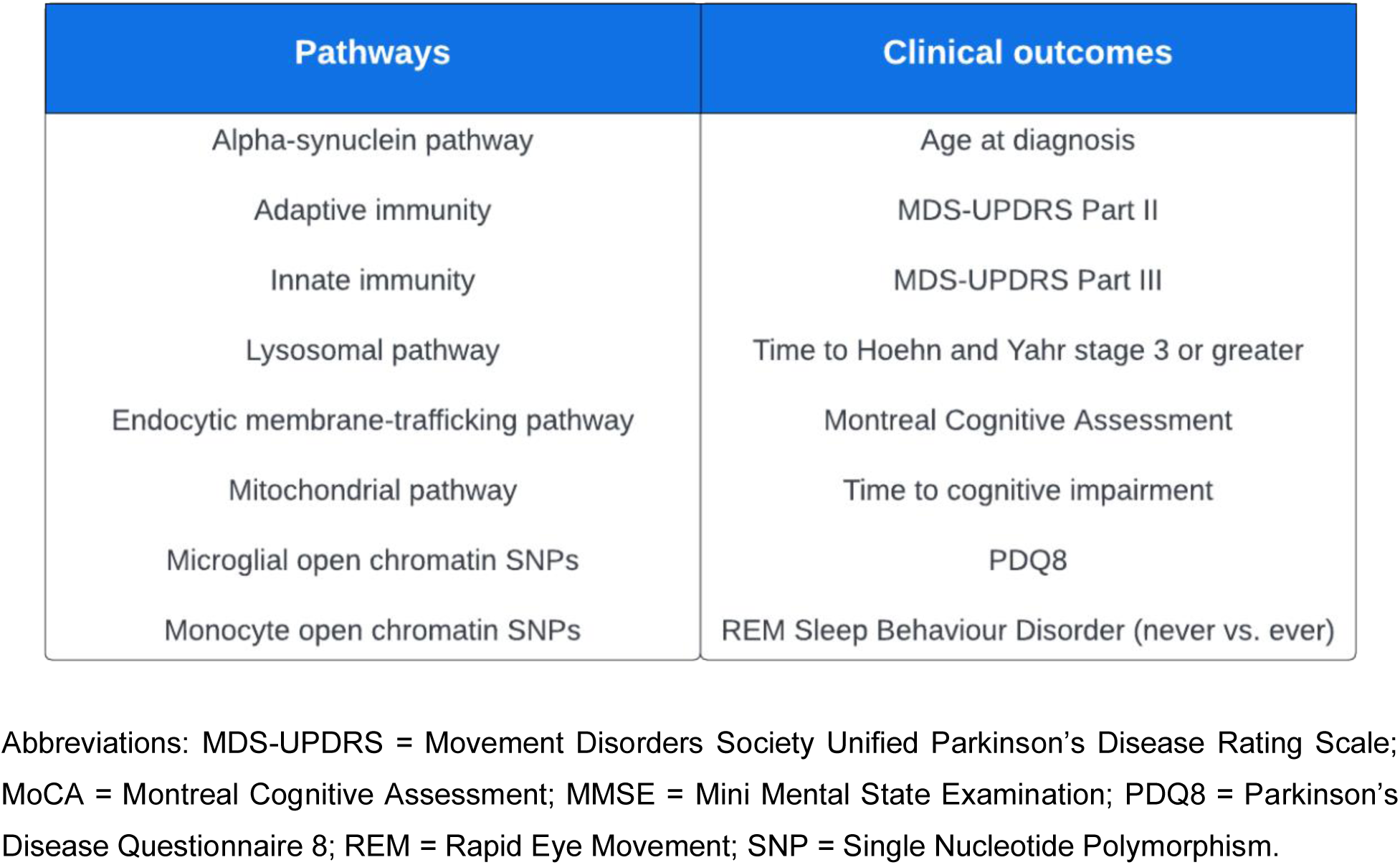
Pathways and clinical outcomes investigated in this study.

We also investigated a selected set of clinical outcomes, based on the assessments that had the most available data across cohorts and that have been frequently used in observational studies as well as clinical trials to measure impairment and progression, such as the Movement Disorders Society Unified Parkinson’s Disease Rating Scale (MDS-UPDRS Part III)^28^ (Figure 1).

## Methods

### In-person cohorts

We meta-analysed data from 18 PD cohorts from Europe and America, including 7,402 participants with PD (Table 1). This included six cohorts in the Accelerating Medicines Partnership (AMP-PD) platform (version 2.5). Key inclusion criteria and cohort features are summarised in the <u>Supplementary Materials</u>. These studies typically recruit patients from specialised hospital clinics, such as movement disorders clinics, and patients are assessed using a combination of clinician assessments (such as the MDS-UPDRS) and self-report questionnaires. We will refer to these as the ‘in-person’ cohorts, in contrast to the Fox Insight study (described below) which is purely online and based on patient-reported outcomes only.

**Table 1.**
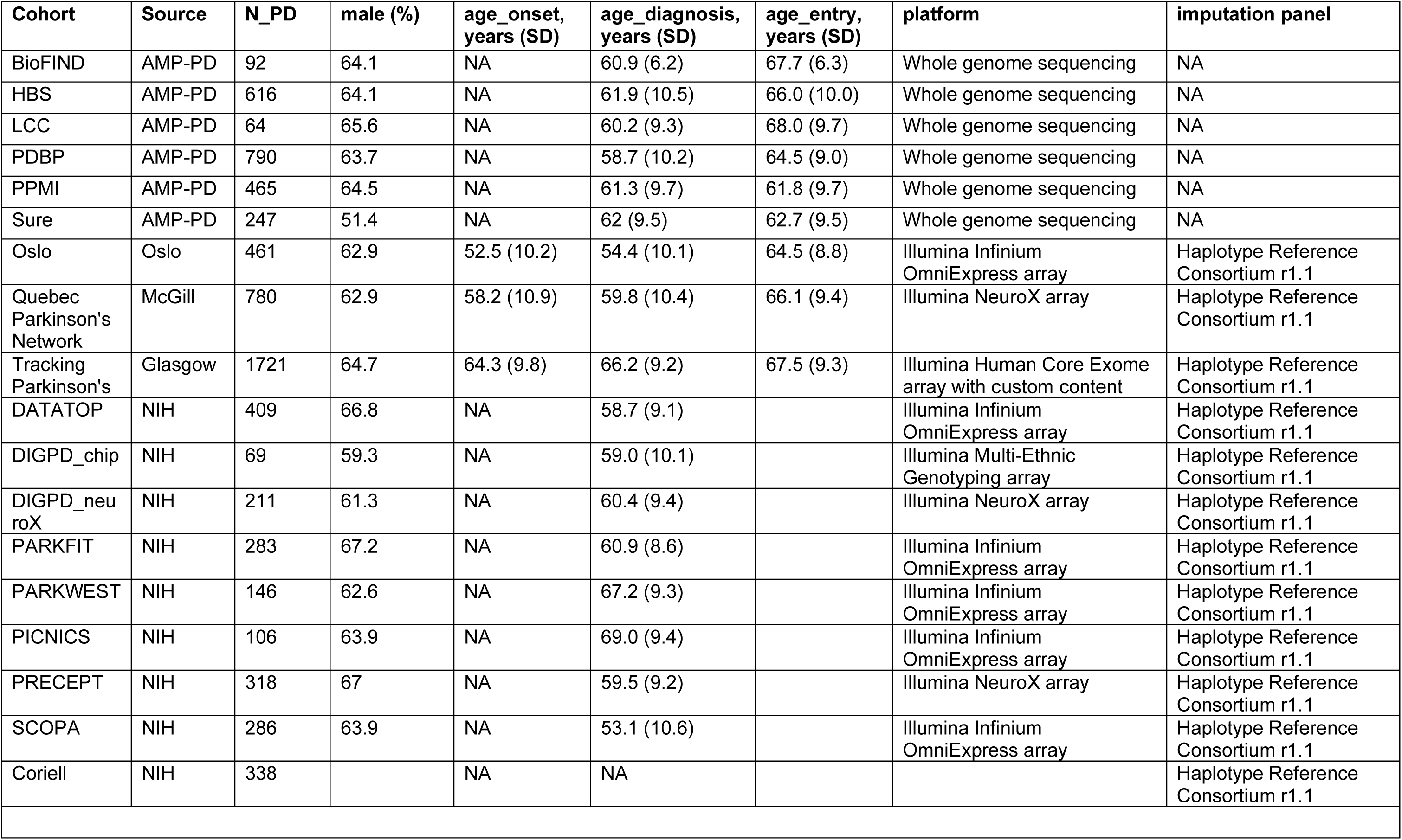
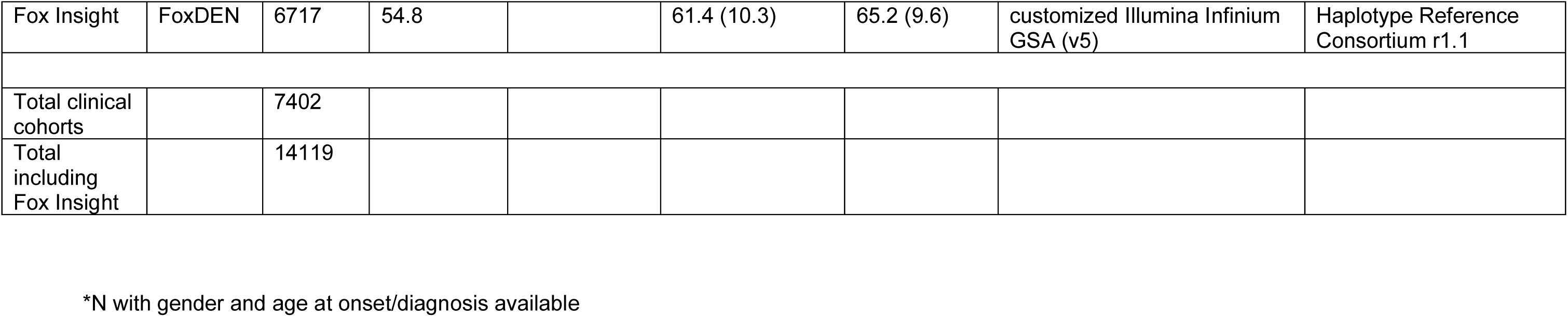
Cohort demographics and genotyping information.

| Cohort | Source | N_PD | male (%) | age_onset,<br>years (SD) | age_diagnosis,<br>years (SD) | age_entry,<br>years (SD) | platform | imputation panel |
| --- | --- | --- | --- | --- | --- | --- | --- | --- |
| BioFIND | AMP-PD | 92 | 64.1 | NA | 60.9 (6.2) | 67.7 (6.3) | Whole genome sequencing | NA |
| HBS | AMP-PD | 616 | 64.1 | NA | 61.9 (10.5) | 66.0 (10.0) | Whole genome sequencing | NA |
| LCC | AMP-PD | 64 | 65.6 | NA | 60.2 (9.3) | 68.0 (9.7) | Whole genome sequencing | NA |
| PDBP | AMP-PD | 790 | 63.7 | NA | 58.7 (10.2) | 64.5 (9.0) | Whole genome sequencing | NA |
| PPMI | AMP-PD | 465 | 64.5 | NA | 61.3 (9.7) | 61.8 (9.7) | Whole genome sequencing | NA |
| Sure | AMP-PD | 247 | 51.4 | NA | 62 (9.5) | 62.7 (9.5) | Whole genome sequencing | NA |
| Oslo | Oslo | 461 | 62.9 | 52.5 (10.2) | 54.4 (10.1) | 64.5 (8.8) | Illumina Infinium<br>OmniExpress array | Haplotype Reference<br>Consortium r1.1 |
| Quebec<br>Parkinson's<br>Network | McGill | 780 | 62.9 | 58.2 (10.9) | 59.8 (10.4) | 66.1 (9.4) | Illumina NeuroX array | Haplotype Reference<br>Consortium r1.1 |
| Tracking<br>Parkinson's | Glasgow | 1721 | 64.7 | 64.3 (9.8) | 66.2 (9.2) | 67.5 (9.3) | Illumina Human Core Exome<br>array with custom content | Haplotype Reference<br>Consortium r1.1 |
| DATATOP | NIH | 409 | 66.8 | NA | 58.7 (9.1) |  | Illumina Infinium<br>OmniExpress array | Haplotype Reference<br>Consortium r1.1 |
| DIGPD_chip | NIH | 69 | 59.3 | NA | 59.0 (10.1) |  | Illumina Multi-Ethnic<br>Genotyping array | Haplotype Reference<br>Consortium r1.1 |
| DIGPD_neu<br>roX | NIH | 211 | 61.3 | NA | 60.4 (9.4) |  | Illumina NeuroX array | Haplotype Reference<br>Consortium r1.1 |
| PARKFIT | NIH | 283 | 67.2 | NA | 60.9 (8.6) |  | Illumina Infinium<br>OmniExpress array | Haplotype Reference<br>Consortium r1.1 |
| PARKWEST | NIH | 146 | 62.6 | NA | 67.2 (9.3) |  | Illumina Infinium<br>OmniExpress array | Haplotype Reference<br>Consortium r1.1 |
| PICNICS | NIH | 106 | 63.9 | NA | 69.0 (9.4) |  | Illumina Infinium<br>OmniExpress array | Haplotype Reference<br>Consortium r1.1 |
| PRECEPT | NIH | 318 | 67 | NA | 59.5 (9.2) |  | Illumina NeuroX array | Haplotype Reference<br>Consortium r1.1 |
| SCOPA | NIH | 286 | 63.9 | NA | 53.1 (10.6) |  | Illumina Infinium<br>OmniExpress array | Haplotype Reference<br>Consortium r1.1 |
| Coriell | NIH | 338 |  | NA | NA |  |  | Haplotype Reference<br>Consortium r1.1 |
| Fox Insight | FoxDEN | 6717 | 54.8 |  | 61.4 (10.3) | 65.2 (9.6) | customized Illumina Infinium GSA (v5) | Haplotype Reference Consortium r1.1 |
| Total clinical cohorts |  | 7402 |  |  |  |  |  |  |
| Total including Fox Insight |  | 14119 |  |  |  |  |  |  |

\*N with gender and age at onset/diagnosis available

### Clinical data

#### Age at diagnosis

Age at onset was not available on AMP-PD so age at diagnosis was used. Although some other cohorts had both age at onset and age at diagnosis available, we analysed just age at diagnosis to maximise consistency across cohorts. If age at diagnosis was missing and age at onset was available, we imputed the age at diagnosis using the mean time from onset to diagnosis in other participants. Age at diagnosis and age at onset are known to be highly correlated (Pearson correlation coefficient ρ =0.92)^4^.

#### Definition of cognitive impairment

Cognitive impairment was defined as a Montreal Cognitive Assessment (MoCA) score of 21 or less or an Mini Mental State Examination (MMSE) score of 26 or less^7^.

#### Definition of RBD

Probable REM Sleep Behaviour Disorder (RBD) was defined from questionnaire data following previous studies. Participants were classified as having RBD if they answered ‘Yes’ on the Mayo Sleep Questionnaire item 1 (‘acting out dreams’)^29–32^. Alternatively, participants were classified as having RBD if they had a total REM Sleep Behaviour Disorder Questionnaire (RBDSQ)^33^ equal to or above 6. Although the original RBDSQ study and others have used a cutoff of 5 points to define RBD^33,34^, other studies show that a higher cutoff of 6 may be more suitable and has higher specificity, particularly in PD patients^35–37^.

#### Missing data

Missing clinical data was not imputed, and total scores (e.g. MDS-UPDRS-III total, PDQ8 total) were only calculated if all items were completed. Clinical outcomes were not standardised so that the resulting effect sizes could be directly interpreted.

Participants were included if they had at least 1 datapoint completed for the outcome of interest through the course of the study. If a participant was missing data for an outcome at all visits, they were excluded from analysis of that outcome.

### Genetic data

Information on the genotyping arrays and imputation reference panel are shown in Table 1. Information about the processing and quality checks performed on the AMP-PD whole genome sequencing data can be found at https://amp-pd.org/whole-genome-data.

Standard quality control of genetic data was conducted within each cohort separately, following standard procedures and using plink v1.9 and v2.0^38,39^. Briefly, we excluded samples with a genotyping rate < 95%, heterozygosity outliers (heterozygosity ≤ −0.15 or ≥ 0.15), and samples whose clinical sex did not match the genetic sex. Non-European ancestry samples were also removed, after clustering with the HapMap 3 data. Related samples were removed (Genome-wide Complex Trait Analysis relatedness from the genetic relatedness matrix > 0.125). We removed variants with genotyping rate < 95%, missingness by haplotype p < 1 x 10^-4^, Hardy-Weinberg Equilibrium p-value < 1 x 10^-4^, and minor allele frequency < 1%.

For the cohorts that were genotyped on SNP arrays, each cohort was imputed to the Haplotype Reference Consortium panel (r1.1) on the Michigan Imputation Server^40^. After imputation of SNP array data, only variants with imputation quality score (R2) > 0.3 were retained for analysis. The AMP-PD genetic data was not imputed as this is whole genome sequencing data. Individuals carrying known PD mutations were not excluded, other than those recruited as part of the genetically-enriched cohorts in AMP-PD.

### Polygenic risk scores

The Polygenic Risk Score (PRS) is a score for each individual based on the weighted sum of risk alleles that individual carries. Allele weights were based on summary statistics from an earlier iteration of the PD GWAS meta-analysis (base dataset)^41^ to avoid overlap between the base and the target datasets.

PRSs were calculated using *PRSice2* v2.3.3 (https://www.prsice.info/)^42,43^. We created pathway-specific PRSs for each pathway of interest. First, the variants/regions for each pathway were extracted from the target dataset (see Supplementary Tables for exact positions). Following the methodology in previous studies^11^, we generated PRSs using variants with a summary statistic p-value of < 0.05 and minor allele frequency > 1%. Linkage disequilibrium (LD) clumping was performed using default settings (r^2^ = 0.1 and distance of 250kb). As we did not have sufficient samples to divide our cohorts into training and testing datasets, we did not perform permutation testing to optimise the p-value threshold and only tested the PRS at p-value threshold 0.05. A previous large-scale study used the same methodology to create pathway-specific PRS and found that these were associated with PD risk^11^. PRSs were standardised within each cohort (each score subtracted from the mean and divided by the standard deviation) prior to analysis.

### Pathway-specific PRS

SNPs were mapped to genes based on physical proximity. The gene sets were based on the curated gene sets representing canonical pathways, publicly available in the Molecular Signatures Database v7.0 (MsigDB)^44,45^. The gene sets and genomic regions for each pathway are provided in Supplementary Tables 1-8. The gene set for the endocytic membrane trafficking pathway was based on Kyoto Encyclopedia of Genes and Genomes (KEGG) data but with additional genes nominated by a literature search conducted by Bandres-Ciga et al.^14^ The gene sets for the mitochondrial pathway were taken from Billingsley et al.^15^, combining both the primary and secondary gene lists. The regions for monocyte and microglia open chromatin were taken from Assay for Transposase-Accessible Chromatin sequencing (ATAC-seq) data^46,47^.

### Statistical analysis

Time to event data (progression to Hoehn and Yahr stage 3+, and significant cognitive impairment) was analysed using Cox proportional hazard models. PD diagnosis was used as the starting time point. The time to event was taken as the first visit where the outcome was met. If the outcome was not met, the time to censoring was taken as the last visit where the outcome was not met. Age at diagnosis, gender, and the first 5 genetic principal components (PCs) were used as covariates.

Longitudinal continuous outcomes (MDS-UPDRS, MoCA, PDQ8) were analysed using linear mixed effects models, adjusting for age at diagnosis, gender, and PC1-PC5. Random effects terms were included for the intercept (to allow for individual variation in baseline scores) and the slope (to allow for individual variation in the rate of progression). In the mixed effects models, we looked at the interaction between the pathway-PRS and years from diagnosis, which is the slope of the model indicating progression in the outcome over time. RBD was analysed as never vs. ever RBD using logistic regression, adjusting for sex, age at diagnosis, and genetic PC1-PC5. Bonferroni correction was applied to adjust for multiple testing for 64 tests (8 pathways x 8 outcomes), corresponding to an adjusted p-value threshold of p < 0.0008.

### Meta-analysis

Results from different in-person cohorts were combined using random-effects meta-analysis in R. Heterogeneity of effects were assessed using Cochran’s Q and I^2^ statistics

### Fox Insight cohort

The Fox Insight study is a large-scale online study of participants in the US. This cohort is uniquely based on patient-reported outcomes only, instead of traditional clinician assessments like the other ‘in-person’ cohorts analysed. For this reason, we did not include the Fox Insight cohort in the main meta-analysis and used it as a separate replication cohort.

Data from the Fox Insight cohort was accessed through the Fox Data Exploration Network (Fox DEN) platform (https://foxden.michaeljfox.org/). We used data from the August 2022 data freeze. Only individuals with an initial diagnosis as well as a current diagnosis of PD were included. A subset of individuals with PD were genotyped by 23andMe on a customised Illumina Infinium GSA (version 5) array. We excluded individuals missing age at diagnosis, sex, or genetic data. The same genetic quality control and imputation was performed as described above.

We created and analysed the pathway-specific PRSs in the Fox Insight cohort in exactly the same manner as described above for the in-person cohorts. We analysed the following clinical outcomes in the Fox Insight cohort: age at diagnosis (as age at onset was not available), MDS-UPDRS Part II, Brief Motor Screen, cognition measured using the PDAQ15, PDQ8, and RBD. Although no clinician assessments were conducted, we selected these outcomes to match as closely to the outcomes analysed in the in-person cohorts.

### Power calculations

We conducted power calculations to determine the effect size that we had sufficient statistical power (>80%) to detect with the current available sample size. We also used power calculation tools to understand and plot the number of samples that would be needed to detect effect sizes that have been previously reported in case-control analyses. Power calculations for generalised linear models (age at onset) were performed using the R package *pwr*. For longitudinal analysis with mixed effects models, we used the R package *simr*^48^. For survival analysis, we used the R package *powerSurvEpi*.

## Results

### Pathway-specific PRS

A total of 7,402 individuals in the in-person cohorts had minimum clinical data (age at diagnosis and sex) and genetic data available across 18 cohorts (Table 1). The number of individuals included in each analysis varied as not all individuals/studies had all clinical outcomes available (see Table 2).

**Table 2.** Results of random-effects meta-analysis for each of the pathway-specific PRS and each clinical outcome.

| Age at diagnosis |  |  |  |  |  |  |  |  |
| --- | --- | --- | --- | --- | --- | --- | --- | --- |
| pathway | beta | se | 95CI | pval | n_studies | I2 | CochransQ_pval | N_inds |
| adaptive_immune | -0.036 | 0.114 | -0.26, 0.19 | 0.752 | 18 | 0.000 | 0.481 | 7402 |
| alpha_synuclein | -0.268 | 0.114 | -0.49, -0.04 | 0.019 | 18 | 0.000 | 0.798 | 7402 |
| innate_immune | 0.029 | 0.115 | -0.20, 0.25 | 0.803 | 18 | 0.000 | 0.836 | 7402 |
| lysosome | -0.303 | 0.114 | -0.53, -0.08 | 0.008 | 18 | 0.000 | 0.856 | 7402 |
| endocytosis | 0.021 | 0.150 | -0.27, 0.31 | 0.891 | 18 | 0.346 | 0.074 | 7402 |
| microglia | -0.231 | 0.128 | -0.48, 0.02 | 0.072 | 18 | 0.168 | 0.253 | 7402 |
| monocytes | -0.261 | 0.114 | -0.48, -0.04 | 0.022 | 18 | 0.000 | 0.521 | 7402 |
| mitochondria | 0.113 | 0.155 | -0.19, 0.42 | 0.469 | 18 | 0.381 | 0.052 | 7402 |
| Cognitive impairment |  |  |  |  |  |  |  |  |
| pathway | beta | se | 95CI | pval | n_studies | I2 | CochransQ_pval | N_inds |
| adaptive_immune | -0.014 | 0.044 | -0.10, 0.07 | 0.745 | 10 | 0.287 | 0.181 | 4804 |
| alpha_synuclein | 0.030 | 0.033 | -0.04, 0.10 | 0.365 | 10 | 0.226 | 0.235 | 4804 |
| innate_immune | -0.021 | 0.044 | -0.11, 0.06 | 0.626 | 10 | 0.190 | 0.268 | 4804 |
| lysosome | 0.016 | 0.034 | -0.05, 0.08 | 0.641 | 10 | 0.134 | 0.320 | 4804 |
| endocytosis | 0.008 | 0.044 | -0.08, 0.09 | 0.847 | 10 | 0.459 | 0.055 | 4804 |
| microglia | 0.016 | 0.034 | -0.05, 0.08 | 0.643 | 10 | 0.000 | 0.842 | 4804 |
| monocytes | 0.028 | 0.033 | -0.04, 0.09 | 0.391 | 10 | 0.075 | 0.373 | 4804 |
| mitochondria | -0.029 | 0.053 | -0.13, 0.08 | 0.583 | 10 | 0.424 | 0.075 | 4804 |
| Hoehn and Yahr stage 3+ |  |  |  |  |  |  |  |  |

| pathway | beta | se | 95CI | pval | n_studies | I2 | CochransQ_pval | N_inds |
| --- | --- | --- | --- | --- | --- | --- | --- | --- |
| adaptive_immune | 0.004 | 0.033 | -0.06, 0.07 | 0.912 | 14 | 0.118 | 0.324 | 5812 |
| alpha_synuclein | 0.021 | 0.027 | -0.03, 0.07 | 0.419 | 14 | 0.352 | 0.094 | 5812 |
| innate_immune | -0.018 | 0.027 | -0.07, 0.04 | 0.516 | 14 | 0.284 | 0.151 | 5812 |
| lysosome | 0.021 | 0.036 | -0.05, 0.09 | 0.560 | 14 | 0.329 | 0.112 | 5812 |
| endocytosis | 0.045 | 0.048 | -0.05, 0.14 | 0.355 | 14 | 0.527 | 0.011 | 5812 |
| microglia | -0.030 | 0.027 | -0.08, 0.02 | 0.266 | 14 | 0.000 | 0.826 | 5812 |
| monocytes | -0.018 | 0.031 | -0.08, 0.04 | 0.572 | 14 | 0.000 | 0.458 | 5812 |
| mitochondria | -0.036 | 0.027 | -0.09, 0.02 | 0.175 | 14 | 0.000 | 0.954 | 5812 |

**MDS-UPDRS Part II**
| pathway | beta | se | 95CI | pval | n_studies | I2 | CochransQ_pval | N_inds |
| --- | --- | --- | --- | --- | --- | --- | --- | --- |
| adaptive_immune | 0.020 | 0.029 | -0.04, 0.08 | 0.500 | 6 | 0.000 | 0.625 | 3498 |
| alpha_synuclein | 0.011 | 0.025 | -0.04, 0.06 | 0.662 | 6 | 0.083 | 0.363 | 3498 |
| innate_immune | -0.013 | 0.025 | -0.06, 0.04 | 0.611 | 6 | 0.000 | 0.670 | 3498 |
| lysosome | -0.013 | 0.027 | -0.07, 0.04 | 0.630 | 6 | 0.000 | 0.664 | 3498 |
| endocytosis | 0.034 | 0.025 | -0.01, 0.08 | 0.171 | 6 | 0.348 | 0.175 | 3498 |
| microglia | -0.023 | 0.025 | -0.07, 0.03 | 0.356 | 6 | 0.416 | 0.128 | 3498 |
| monocytes | -0.024 | 0.025 | -0.07, 0.03 | 0.340 | 6 | 0.109 | 0.346 | 3498 |
| mitochondria | -0.003 | 0.025 | -0.05, 0.05 | 0.906 | 6 | 0.000 | 0.872 | 3498 |

**MDS-UPDRS Part III**
| pathway | beta | se | 95CI | pval | n_studies | I2 | CochransQ_pval | N_inds |
| --- | --- | --- | --- | --- | --- | --- | --- | --- |
| adaptive_immune | 0.079 | 0.068 | -0.05, 0.21 | 0.246 | 8 | 0.323 | 0.170 | 3933 |
| alpha_synuclein | 0.075 | 0.047 | -0.02, 0.17 | 0.105 | 8 | 0.000 | 0.860 | 3933 |
| innate_immune | 0.016 | 0.069 | -0.12, 0.15 | 0.811 | 8 | 0.470 | 0.067 | 3933 |
| lysosome | 0.022 | 0.047 | -0.07, 0.11 | 0.637 | 8 | 0.000 | 0.466 | 3933 |
| endocytosis | -0.010 | 0.057 | -0.12, 0.10 | 0.857 | 8 | 0.250 | 0.229 | 3933 |
| microglia | -0.071 | 0.046 | -0.16, 0.02 | 0.122 | 8 | 0.103 | 0.350 | 3933 |
| monocytes | -0.059 | 0.046 | -0.15, 0.03 | 0.203 | 8 | 0.154 | 0.309 | 3933 |
| mitochondria | -0.002 | 0.047 | -0.09, 0.09 | 0.971 | 8 | 0.000 | 0.826 | 3933 |

| **Montreal Cognitive Assessment** |  |  |  |  |  |  |  |  |
| --- | --- | --- | --- | --- | --- | --- | --- | --- |
| pathway | beta | se | 95CI | pval | n_studies | I2 | CochransQ_pval | N_inds |
| adaptive_immune | -0.014 | 0.015 | -0.04, 0.01 | 0.345 | 7 | 0.000 | 0.637 | 3685 |
| alpha_synuclein | -0.002 | 0.015 | -0.03, 0.03 | 0.878 | 7 | 0.000 | 0.565 | 3685 |
| innate_immune | 0.009 | 0.017 | -0.02, 0.04 | 0.608 | 7 | 0.222 | 0.260 | 3685 |
| lysosome | -0.003 | 0.016 | -0.03, 0.03 | 0.857 | 7 | 0.555 | 0.036 | 3685 |
| endocytosis | -0.007 | 0.013 | -0.03, 0.02 | 0.591 | 7 | 0.462 | 0.084 | 3685 |
| microglia | -0.004 | 0.013 | -0.03, 0.02 | 0.768 | 7 | 0.000 | 0.650 | 3685 |
| monocytes | -0.008 | 0.013 | -0.03, 0.02 | 0.563 | 7 | 0.115 | 0.341 | 3685 |
| mitochondria | -0.012 | 0.019 | -0.05, 0.02 | 0.507 | 7 | 0.350 | 0.161 | 3685 |

| **PDQ8** |  |  |  |  |  |  |  |  |
| --- | --- | --- | --- | --- | --- | --- | --- | --- |
| pathway | beta | se | 95CI | pval | n_studies | I2 | CochransQ_pval | N_inds |
| adaptive_immune | -0.006 | 0.042 | -0.09, 0.08 | 0.885 | 3 | 0.778 | 0.011 | 2755 |
| alpha_synuclein | 0.012 | 0.030 | -0.05, 0.07 | 0.685 | 3 | 0.478 | 0.147 | 2755 |
| innate_immune | 0.001 | 0.019 | -0.04, 0.04 | 0.969 | 3 | 0.000 | 0.795 | 2755 |
| lysosome | -0.006 | 0.020 | -0.04, 0.03 | 0.773 | 3 | 0.000 | 0.943 | 2755 |
| endocytosis | -0.002 | 0.020 | -0.04, 0.04 | 0.924 | 3 | 0.000 | 0.837 | 2755 |

| microglia | 0.043 | 0.062 | -0.08, 0.16 | 0.487 | 3 | 0.803 | 0.006 | 2755 |
| --- | --- | --- | --- | --- | --- | --- | --- | --- |
| monocytes | 0.030 | 0.052 | -0.07, 0.13 | 0.570 | 3 | 0.750 | 0.018 | 2755 |
| mitochondria | 0.061 | 0.053 | -0.04, 0.17 | 0.247 | 3 | 0.755 | 0.017 | 2755 |
| **RBD (never vs. ever)** |  |  |  |  |  |  |  |  |
| pathway | beta | se | 95CI | pval | n_studies | I2 | CochransQ_pval | N_inds |
| adaptive_immune | 0.002 | 0.036 | -0.07, 0.07 | 0.963 | 9 | 0.571 | 0.017 | 4414 |
| alpha_synuclein | -0.012 | 0.016 | -0.04, 0.02 | 0.431 | 9 | 0.000 | 0.450 | 4414 |
| innate_immune | 0.034 | 0.057 | -0.08, 0.15 | 0.556 | 9 | 0.631 | 0.006 | 4414 |
| lysosome | 0.010 | 0.016 | -0.02, 0.04 | 0.527 | 9 | 0.000 | 0.563 | 4414 |
| endocytosis | 0.005 | 0.016 | -0.03, 0.04 | 0.756 | 9 | 0.000 | 0.558 | 4414 |
| microglia | -0.002 | 0.018 | -0.04, 0.03 | 0.919 | 9 | 0.234 | 0.235 | 4414 |
| monocytes | 0.012 | 0.016 | -0.02, 0.04 | 0.425 | 9 | 0.000 | 0.848 | 4414 |
| mitochondria | 0.020 | 0.016 | -0.01, 0.05 | 0.211 | 9 | 0.000 | 0.883 | 4414 |

We did not find that any of the 8 pathway-specific PRSs were significantly associated with any of the 8 clinical outcomes of interest (Table 2) (Bonferroni-adjusted p-value threshold < 0.0008). Forest plots showing effect estimates on a cohort level for each of the outcomes and each of the pathways are available through our online browser (https://manuelatan.shinyapps.io/pathway-prs-app/).

There were only a few nominal associations (p < 0.05) for age at diagnosis (highlighted in Table 2). Here we found that the alpha-synuclein pathway, lysosomal pathway, and monocytes open chromatin regions showed a trend towards association with lower age at diagnosis (Figure 1). However these did not pass correction for multiple testing.

We also checked whether the pathway-specific PRSs were correlated with each other. In the largest cohort, PROBAND, we showed that the PRSs were not highly correlated with one another, except for the monocytes and microglia open-chromatin region PRSs (r = 0.70, p < 2 x 10^-16^) and the endocytosis and adaptive immune PRSs (r = 0.59, p < 2 x 10^-16^) (Figure 2).

**Figure 2.**
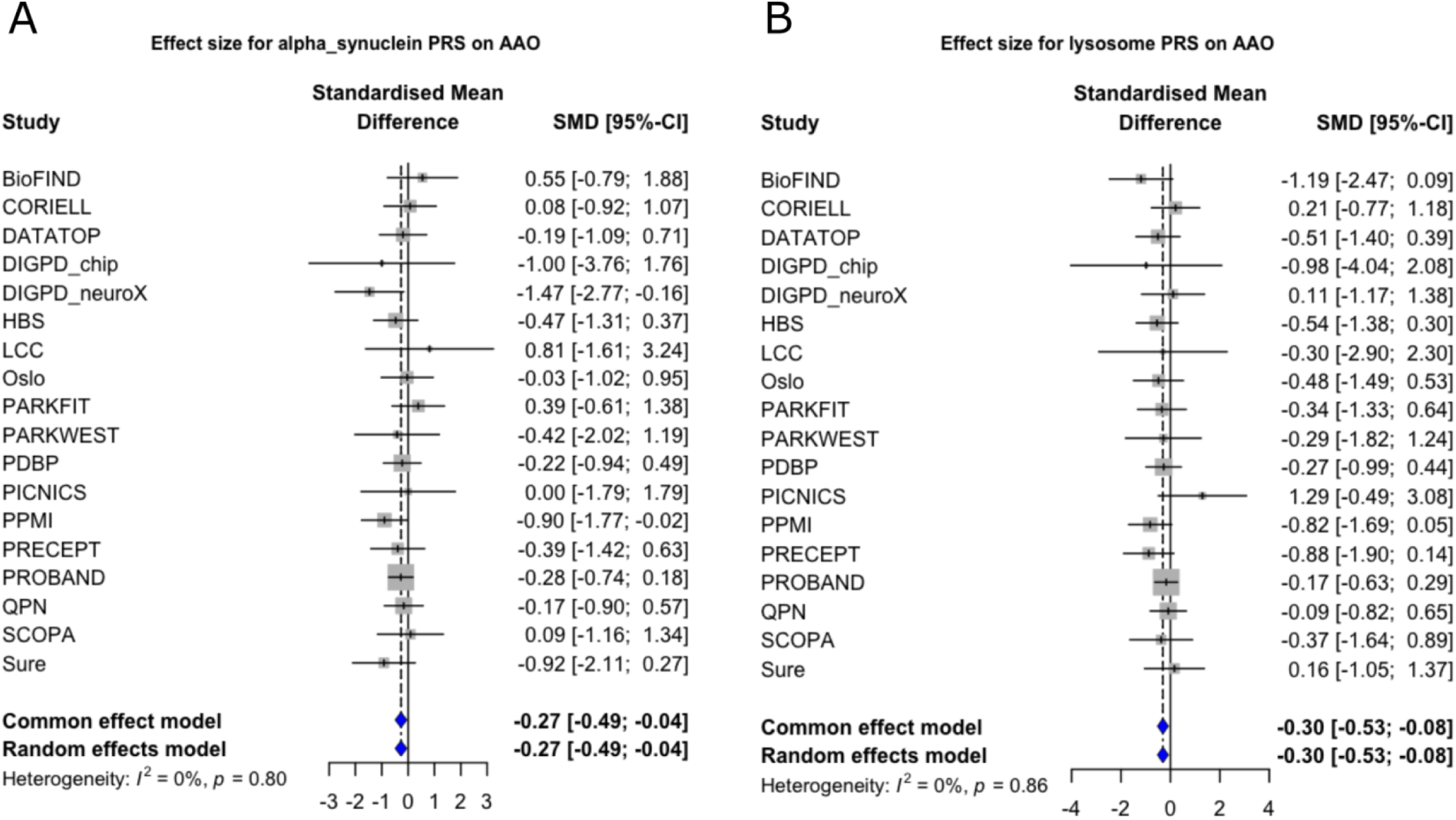
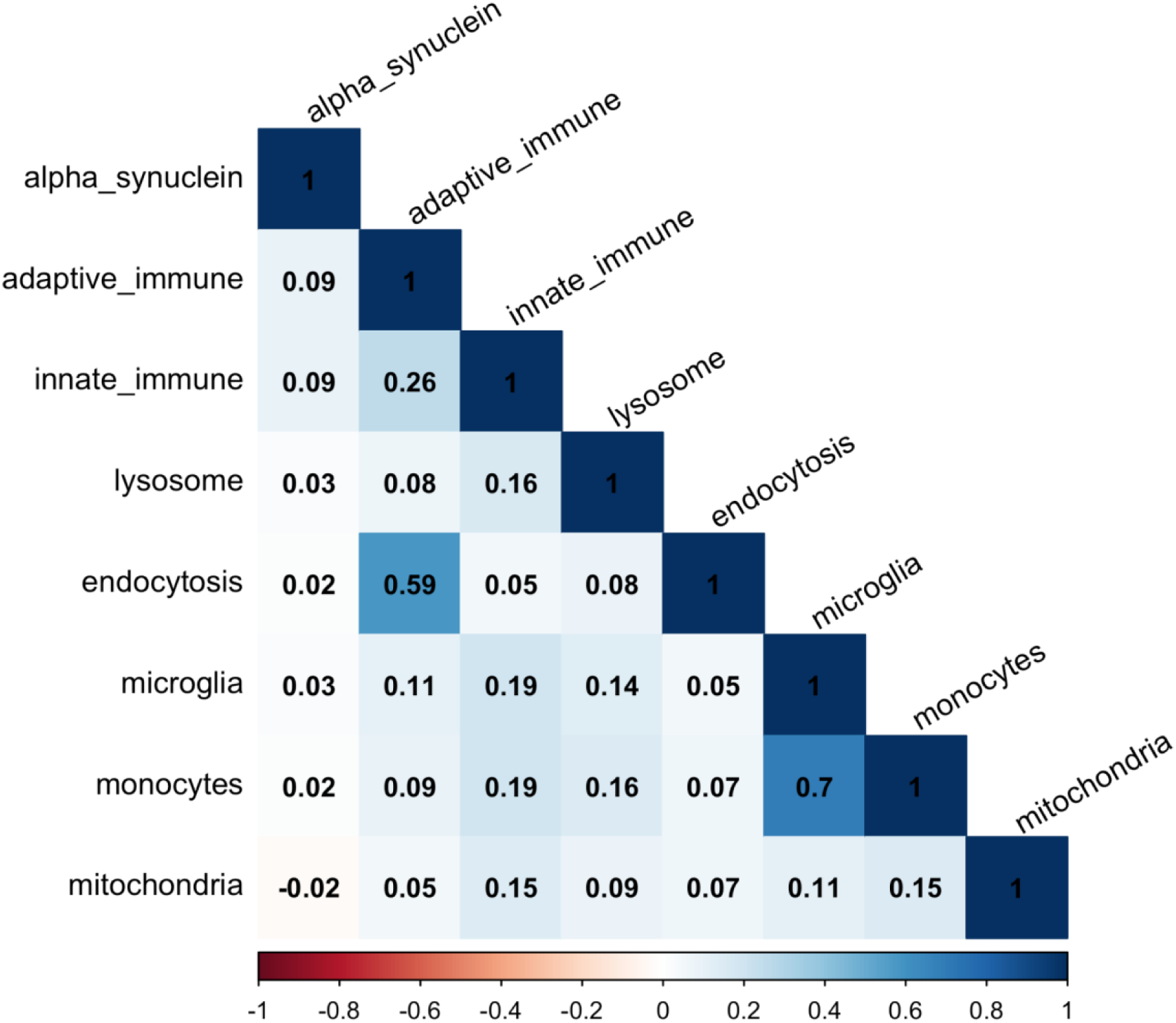
Correlation between pathway-specific PRSs in the largest cohort, Tracking Parkinson’s (PROBAND) Forest plots for A) alpha-synuclein PRS on age at onset; B) lysosomal PRS on age at onset. AAO = age at onset; CI = Confidence Interval; PRS = Polygenic Risk Score; SMD = standardised mean difference.

### Fox Insight

A total of 6,717 individuals in the Fox Insight cohort had minimum clinical data and genetic data available. The number of individuals included in each analysis is shown in Table 3. We did not find that any of the 8 pathway-specific PRSs were significantly associated with any of the clinical outcomes surpassing Bonferroni correction (p < 0.0008).

**Table 3.** Results for pathway-specific PRS in the Fox Insight cohort.

| **Age at onset** |  |  |  |  |
| --- | --- | --- | --- | --- |
| pathway | beta | se | Pvalue | N_inds |
| adaptive_immune | -0.106 | 0.125 | 0.398 | 6717 |
| alpha_synuclein | -0.147 | 0.125 | 0.241 | 6717 |
| innate_immune | -0.049 | 0.125 | 0.696 | 6717 |
| lysosome | 0.148 | 0.125 | 0.237 | 6717 |
| endocytosis | -0.250 | 0.125 | 0.045 | 6717 |
| microglia | -0.107 | 0.125 | 0.393 | 6717 |
| monocytes | -0.132 | 0.125 | 0.293 | 6717 |
| mitochondria | 0.081 | 0.125 | 0.520 | 6717 |

| **MDS-UPDRS Part II** |  |  |  |  |  |
| --- | --- | --- | --- | --- | --- |
| pathway | beta | se | Pvalue | N_obs | N_inds |
| adaptive_immune | -0.045 | 0.031 | 0.143 | 27685 | 6685 |
| alpha_synuclein | -0.067 | 0.031 | 0.032 | 27685 | 6685 |
| innate_immune | 0.001 | 0.031 | 0.986 | 27685 | 6685 |
| lysosome | -0.032 | 0.031 | 0.308 | 27685 | 6685 |
| endocytosis | 0.029 | 0.031 | 0.353 | 27685 | 6685 |
| microglia | -0.060 | 0.031 | 0.053 | 27685 | 6685 |
| monocytes | -0.019 | 0.031 | 0.553 | 27685 | 6685 |
| mitochondria | 0.017 | 0.031 | 0.576 | 27685 | 6685 |

| **Brief Motor Screen** |  |  |  |  |  |
| --- | --- | --- | --- | --- | --- |
| pathway | beta | se | Pvalue | N_obs | N_inds |
| adaptive_immune | 0.001 | 0.018 | 0.939 | 21144 | 4585 |
| alpha_synuclein | 0.015 | 0.018 | 0.389 | 21144 | 4585 |
| innate_immune | 0.004 | 0.018 | 0.816 | 21144 | 4585 |
| lysosome | 0.018 | 0.018 | 0.298 | 21144 | 4585 |
| endocytosis | 0.031 | 0.018 | 0.087 | 21144 | 4585 |
| microglia | -0.005 | 0.018 | 0.757 | 21144 | 4585 |
| monocytes | -0.011 | 0.018 | 0.521 | 21144 | 4585 |
| mitochondria | -0.006 | 0.018 | 0.758 | 21144 | 4585 |

| **Cognition and daily activities (PDAQ15)** |  |  |  |  |  |
| --- | --- | --- | --- | --- | --- |
| pathway | beta | se | Pvalue | N_obs | N_inds |
| adaptive_immune | -0.021 | 0.033 | 0.539 | 27393 | 6696 |
| alpha_synuclein | 0.005 | 0.034 | 0.889 | 27393 | 6696 |
| innate_immune | -0.020 | 0.034 | 0.558 | 27393 | 6696 |
| lysosome | -0.050 | 0.033 | 0.134 | 27393 | 6696 |
| endocytosis | -0.017 | 0.034 | 0.610 | 27393 | 6696 |
| microglia | -0.003 | 0.033 | 0.931 | 27393 | 6696 |
| monocytes | 0.000 | 0.034 | 0.989 | 27393 | 6696 |
| mitochondria | -0.041 | 0.034 | 0.227 | 27393 | 6696 |

| **PDQ8** |  |  |  |  |  |
| --- | --- | --- | --- | --- | --- |
| pathway | beta | se | Pvalue | N_obs | N_inds |
| adaptive_immune | -0.015 | 0.018 | 0.395 | 49072 | 6716 |
| alpha_synuclein | -0.023 | 0.018 | 0.198 | 49072 | 6716 |
| innate_immune | -0.027 | 0.018 | 0.139 | 49072 | 6716 |
| lysosome | -0.005 | 0.018 | 0.793 | 49072 | 6716 |
| endocytosis | -0.026 | 0.018 | 0.158 | 49072 | 6716 |
| microglia | -0.031 | 0.018 | 0.080 | 49072 | 6716 |
| monocytes | -0.015 | 0.018 | 0.397 | 49072 | 6716 |
| mitochondria | -0.009 | 0.018 | 0.630 | 49072 | 6716 |

| **RBD (never vs. ever)** |  |  |  |  |
| --- | --- | --- | --- | --- |
| pathway | beta | se | Pvalue | N_inds |
| adaptive_immune | -0.004 | 0.006 | 0.541 | 6669 |
| alpha_synuclein | -0.011 | 0.006 | 0.068 | 6669 |
| innate_immune | -0.007 | 0.006 | 0.275 | 6669 |
| lysosome | -0.002 | 0.006 | 0.736 | 6669 |
| endocytosis | -0.018 | 0.006 | 0.003 | 6669 |
| microglia | -0.012 | 0.006 | 0.053 | 6669 |
| monocytes | -0.018 | 0.006 | 0.003 | 6669 |
| mitochondria | -0.003 | 0.006 | 0.590 | 6669 |

### Power calculations

To support the interpretation of our negative results, we performed a number of power analyses. We first estimated the theoretical effect sizes that would have been required for our study to have 80% power to detect a true association at a Bonferroni-corrected significance level of p < 0.0008. In the survival analysis for H&Y progression to stage 3 (N = 5,812), we calculate that the hazard ratio (effect size) would need to be at least 1.09. To compare this to previously reported effect sizes in PD risk, we looked at the largest pathway PRS study of PD risk^11^. In that study, the largest reported effect size (Odds Ratio) for the pathways we examined was 1.21 for the adaptive immune pathway in the discovery phase but only 1.08 in the replication cohort. Alternatively, we would need to include over 16,000 participants to detect an effect with hazard ratio 1.05.

In logistic regression analysis of RBD (N = 4,414), we estimate that an effect size of OR = 1.03 (Cohen’s effect size f^2^ = 0.0065) would be required for 80% power. In comparison, previous genetic analyses of probable RBD in PD have shown ORs for top genetic risk loci up between 1.18 and 2.84 with a sample size of 2,843 cases and 139,636 controls^49^.

Next we calculated the sample sizes that would be required for 80% power given an effect size equivalent to the largest single-cohort effects observed in our meta-analysis. For the age at onset linear model, assuming a Cohen’s effect size f^2^ = 0.0008 as estimated in the Tracking Parkinson’s cohort, we estimate that we had 18.6% power with our existing sample size of 14,119 individuals (across both clinical cohorts and the Fox Insight cohort). To achieve 80% power with the same effect size, we would need 34,205 samples.

For survival analysis, we estimated that we had 20.8% power in the survival analysis for progression to H&Y3+. For this calculation, we used our existing sample size of 5,812 individuals, a postulated hazard ratio of 1.05, variance of 1 as the PRSs were standardised, proportion of subjects who met the outcome of interest 0.47 (based on the Oslo cohort), correlation coefficient of 0.065 between the covariate of interest (PRS) and other covariates (age at diagnosis) based on the Oslo cohort, and type 1 error rate/ alpha of 0.00078 (0.05/64).

It is important to note that these power calculations are limited as they are assumed to be within a single cohort, whereas meta-analysis of multiple cohorts with variation in their effect sizes and other factors is likely to reduce power further.

### Progression PRS

We conducted some exploratory analyses to see whether PRSs created from PD phenotypes/progression GWAS, instead of case-control GWAS, were associated with clinical outcomes. Due to limited power we did not attempt to create pathway-stratified PRSs for this analysis.

#### Age at onset PRS

We created an age-at-onset PRS from the PD age at onset GWAS^4^, using p-value thresholds of 5 x 10^-8^, 1 x 10^-5^, and 0.05. As most of our cohorts were included in the initial GWAS, we only tested the PRS in the Fox Insight cohort to avoid sample overlap (N = 6,717). We applied Bonferroni correction for the number of tests within this phenotype (0.05/3 = 0.017). We found that the PRS was associated with age at diagnosis in the Fox Insight cohort at the most liberal p-value threshold 0.05, with higher PRSs associated with higher age at onset (beta = 0.31, p = 0.014) (Table 4). There was also a trend for the other PRSs at other p-value thresholds to be associated with age at onset though these were not significant (Table 4).

**Table 4.**
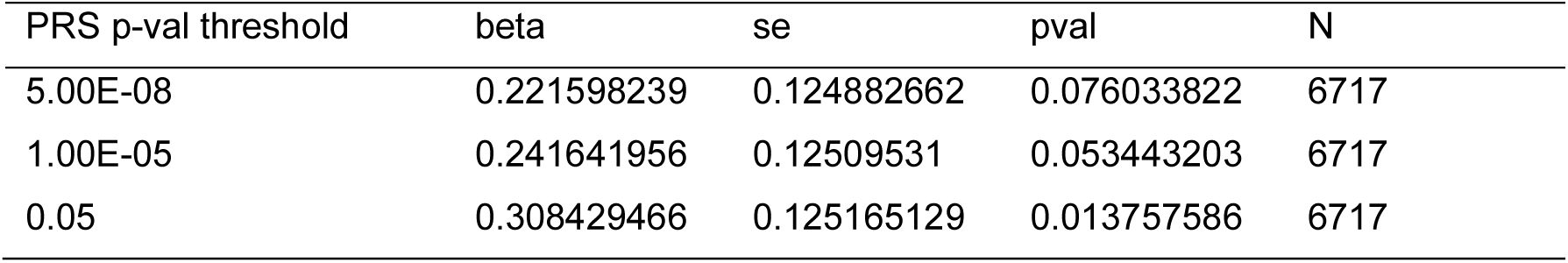
Age at onset PRS at different p-value thresholds tested against age at onset in the Fox Insight cohort.

#### H&Y PRS

We created a PRS for progression to H&Y3+ from a recent progression GWAS^50^, using p-value thresholds of 5 x 10^-8^, 1 x 10^-5^, and 0.05. To avoid sample overlap with the original GWAS, we tested the PRS in the AMP-PD cohorts (excluding PPMI as this was included in the original GWAS) and the Quebec Parkinson’s Network. Hoehn and Yahr stage data was not available in the Fox Insight cohort as this was an online-only study. In a random-effects meta-analysis, we did not find that the PRS at any of the three p-value thresholds was associated with progression to H&Y3+ in the testing cohorts (Supplementary Table 9).

### Public browser

We have made our results publicly available with an online browser https://manuelatan.shinyapps.io/pathway-prs-app/ so that others can view results and forest-plots, download summary statistics and meta-analyse with their own data.

## Discussion

We show clearly that none of the 8 pathway-specific PRSs based on PD risk were associated with clinical outcomes or progression in PD. This is the largest study to date that has looked at the association between PRSs and clinical outcomes in PD, and we replicated these null findings in the large-scale online study of patient-reported outcomes, Fox Insight.

Power analyses indicate that albeit underpowered to detect very small effects of pathway-specific genetic burden on clinical PD outcomes, our study was in theory large enough to capture moderate effects sizes that could reasonably be hypothesized based on previous results. Our findings may therefore suggest that the genetic architecture of sporadic PD risk has limited impact on outcomes or progression once an individual has developed PD. This notion is to some degree also supported by the large-scale GWASs of PD phenotypes. The PD age at onset GWAS^4^ identified some loci that were also PD risk loci (including *SNCA, TMEM175/GASK)* but other PD risk loci clearly did not affect PD age at onset (such as *GCH1, MAPT*). Other large-scale GWASs of PD progression outcomes, such as motor and cognitive progression, dementia, and mortality have identified progression loci that are largely distinct from the PD risk loci, with the exception of GBA1^7,9,10,26,50^.

Our findings are also consistent with explorative analyses of clinical outcomes presented in previous PRS studies of PD. Although multiple studies have confirmed that various pathway-specific PRSs are associated with PD risk^11,14^, several of the same studies have shown that the PRSs were not associated with age at onset of PD^14,15^. Other studies have shown contradictory findings. Lüth et al.^51^ showed that the mitochondrial PRS was associated with earlier age at PD onset, in the opposite direction to Billingsley et al.^15^ and Dehestani et al.^52^ where the mitochondrial PRS was associated with later age at onset. However in general many of these studies looking at clinical outcomes have been exploratory and performed multiple tests. Only one previous study has looked at the PRSs in relation to progression of the UPDRS III and MoCA, and did not find that either mitochondrial PRS or autophagy-lysosomal PRS were associated with progression of clinical scores^52^, in line with our current findings.

Importantly, our ability to demonstrate associations between pathway-specific genetic risk and clinical outcomes is limited not only by statistical power, but also by a number of methodological challenges and caveats. Defining biological pathways is complicated, and the gene lists or genomic annotations used in our study were based on previous publications and may not have been optimal. Systematic exploration of different approaches to pathway curation was beyond the scope of the present study, but should be pursued in future work. If gene lists become more representative of actual pathogenic networks, they are also likely to generate more sensitive pathway-specific PRS. Likewise, the assignment of SNPs to genes was based merely on genomic position, which we acknowledge is a crude approach with a number of caveats. Future efforts taking advantage of e.g. eQTL data to link SNPs to genes may improve PRS sensitivity further.

Furthermore, the major clinical outcome measures used in the present study mainly capture broad aspects of disease progression, and may have highly heterogeneous pathogenic underpinnings on the molecular level. It is therefore possible that pathway-specific PRSs reflect biomarkers and pathology markers more accurately than clinical outcomes. We showed in a separate study that the PD lysosomal PRS was associated with Lewy body pathology in ‘pure’ Lewy body disease patients without Alzheimer’s disease co-pathology^53^. Following these results, we next demonstrated that lysosomal PD-PRS is also associated with cognitive progression in a similar subgroup of PD patients with low AD-risk^54^, illustrating how cognitive decline is likely driven by multiple independent mechanisms. More advanced study designs may therefore be required to disentangle causal complexities in order to successfully link genetic risk profiles to both molecular disease mechanisms and clinical outcomes in PD.

We conducted some exploratory analyses to see whether PRSs created from PD phenotype GWASs, such as age at onset and progression, instead of PD risk, were associated with clinical outcomes. There was some evidence that PRSs created from the PD age at onset GWAS^4^ were associated with age at onset in independent cohorts. We did not find that PRSs created from the PD H&Y3 GWAS were associated with progression to H&Y3 in independent cohorts^50^. However these analyses were exploratory and not the main focus of this article. The sample sizes of the PD phenotype GWASs as well as the testing cohorts are still very small compared to the PD risk GWASs, due to the challenges in systematic collection and harmonisation of detailed longitudinal clinical data across many cohorts. In addition the measurement of clinical phenotypes and progression are affected by many factors such as measurement error and medication effects, so the phenotypes are far ‘noisier’ than case-control studies. This makes the identification and robust replication of loci for PD progression extremely challenging.

This study has several limitations. Our main limitation is in statistical power and sample sizes. However this is the largest study to date to examine the relationship between PD PRS and clinical outcomes or progression. We highlight that it takes many years and intensive resources to collect this type of detailed clinical data through longitudinal observational studies, as well as to harmonise clinical data across cohorts. It is important for funders to direct more resources into generating this type of data in different ancestry groups and make the data easily accessible to researchers.

An additional limitation is that this study was conducted only in samples of European ancestry and PRSs developed in European ancestries have limited predictive performance in other ancestry groups^55^. We did not have access to detailed clinical data from participants of other ancestries, however efforts are already underway to collate and share this data (https://gp2.org/).

Thus we believe further investigation of pathway-specific PRSs is warranted, also investigating PRSs from genome-wide progression studies. This work will help us to better understand the biology of PD progression and predict clinical progression.

## Data Availability

Web browser with summary results: https://manuelatan.shinyapps.io/pathway-prs-app/
AMP-PD data is available on application: https://amp-pd.org/.
Tracking Parkinson's data is available through the Tracking Parkinson's portal: https://www.trackingparkinsons.org.uk/about-1/data/. All other individual level data was provided on request to the individual principal investigators.

https://manuelatan.shinyapps.io/pathway-prs-app/

## Data availability

Web browser with summary results: https://manuelatan.shinyapps.io/pathway-prs-app/

AMP-PD data is available on application: https://amp-pd.org/.

Tracking Parkinson’s data is available through the Tracking Parkinson’s portal: https://www.trackingparkinsons.org.uk/about-1/data/. All other individual level data was provided on request to the individual principal investigators.

## Code availability

GitHub link

NIH analysis: https://github.com/hirotaka-i/prs_progression

Main funding sources: Southern and Eastern Norway Regional Health Authority, Michael J Fox Foundation for Parkinson’s Research

The Tracking Parkinson’s (PRoBaND) cohort (grant reference J-1101) was funded by Parkinson’s UK.

The Parkinsonism: Incidence, Cognition and Non-motor heterogeneity in Cambridgeshire (PICNICS) study was funded by the Cure Parkinson’s Trust, the Van Geest Foundation, the Medical Research Council, Parkinson’s UK, and the NIHR Cambridge Biomedical Research Centre (NIHR203312).The views expressed are those of the authors and not necessarily those of the NHS, the NIHR or the Department of Health.

## AMP PD Acknowledgement

Data used in the preparation of this article were obtained from the Accelerating Medicine Partnership® (AMP®) Parkinson’s Disease (AMP PD) Knowledge Platform. For up-to-date information on the study, visit https://www.amp-pd.org.

The AMP® PD program is a public-private partnership managed by the Foundation for the National Institutes of Health and funded by the National Institute of Neurological Disorders and Stroke (NINDS) in partnership with the Aligning Science Across Parkinson’s (ASAP) initiative; Celgene Corporation, a subsidiary of Bristol-Myers Squibb Company; GlaxoSmithKline plc (GSK); The Michael J. Fox Foundation for Parkinson’s Research; AbbVie Inc.; Pfizer Inc.; Sanofi US Services Inc.; and Verily Life Sciences.

ACCELERATING MEDICINES PARTNERSHIP and AMP are registered service marks of the U.S. Department of Health and Human Services.

## AMP PD Cohort Acknowledgements

Clinical data and biosamples used in preparation of this article were obtained from the (i) Michael J. Fox Foundation for Parkinson’s Research (MJFF) and National Institutes of Neurological Disorders and Stroke (NINDS) BioFIND study, (ii) Harvard Biomarkers Study (HBS), (iii) National Institute on Aging (NIA) International Lewy Body Dementia Genetics Consortium Genome Sequencing in Lewy Body Dementia Case-control Cohort (LBD), (iv) MJFF LRRK2 Cohort Consortium (LCC), (v) NINDS Parkinson’s Disease Biomarkers Program (PDBP), (vi) MJFF Parkinson’s Progression Markers Initiative (PPMI), and (vii) NINDS Study of Isradipine as a Disease-modifying Agent in Subjects With Early Parkinson Disease, Phase 3 (STEADY-PD3) and (viii) the NINDS Study of Urate Elevation in Parkinson’s Disease, Phase 3 (SURE-PD3).

BioFIND is sponsored by The Michael J. Fox Foundation for Parkinson’s Research (MJFF) with support from the National Institute for Neurological Disorders and Stroke (NINDS). The BioFIND Investigators have not participated in reviewing the data analysis or content of the manuscript. For up-to-date information on the study, visit michaeljfox.org/biofind.

Genome sequence data for the Lewy body dementia case-control cohort were generated at the Intramural Research Program of the U.S. National Institutes of Health. The study was supported in part by the National Institute on Aging (program #: 1ZIAAG000935) and the National Institute of Neurological Disorders and Stroke (program #: 1ZIANS003154).

The Harvard Biomarker Study (HBS) is a collaboration of HBS investigators [full list of HBS investigators found at https://www.bwhparkinsoncenter.org/biobank/] and funded through philanthropy and NIH and Non-NIH funding sources. The HBS Investigators have not participated in reviewing the data analysis or content of the manuscript.

Data used in preparation of this article were obtained from The Michael J. Fox Foundation sponsored LRRK2 Cohort Consortium (LCC). The LCC Investigators have not participated in reviewing the data analysis or content of the manuscript. For up-to-date information on the study, visit https://www.michaeljfox.org/biospecimens).

PPMI is sponsored by The Michael J. Fox Foundation for Parkinson’s Research and supported by a consortium of scientific partners: [list the full names of all of the PPMI funding partners found at https://www.ppmi-info.org/about-ppmi/who-we-are/study-sponsors]. The PPMI investigators have not participated in reviewing the data analysis or content of the manuscript. For up-to-date information on the study, visit www.ppmi-info.org.

The Parkinson’s Disease Biomarker Program (PDBP) consortium is supported by the National Institute of Neurological Disorders and Stroke (NINDS) at the National Institutes of Health. A full list of PDBP investigators can be found at https://pdbp.ninds.nih.gov/policy.

The PDBP investigators have not participated in reviewing the data analysis or content of the manuscript.

The Study of Isradipine as a Disease-modifying Agent in Subjects With Early Parkinson Disease, Phase 3 (STEADY-PD3) is funded by the National Institute of Neurological Disorders and Stroke (NINDS) at the National Institutes of Health with support from The Michael J. Fox Foundation and the Parkinson Study Group. For additional study information, visit https://clinicaltrials.gov/ct2/show/study/NCT02168842. The STEADY-PD3 investigators have not participated in reviewing the data analysis or content of the manuscript.

The Study of Urate Elevation in Parkinson’s Disease, Phase 3 (SURE-PD3) is funded by the National Institute of Neurological Disorders and Stroke (NINDS) at the National Institutes of Health with support from The Michael J. Fox Foundation and the Parkinson Study Group. For additional study information, visit https://clinicaltrials.gov/ct2/show/NCT02642393. The SURE-PD3 investigators have not participated in reviewing the data analysis or content of the manuscript.

## Ethics

For all cohorts on the AMP-PD platform, prior to releasing de-identified data, the AMP PD program receives the following written assurances from the cohorts contributing data to the AMP PD Knowledge Platform regarding the shareability of their participant data:

- Participant is fully consented for broad data sharing in a manner consistent with applicable laws enabling sharing with AMP PD users
- Participant is consented for broad individual level clinical and genomics data sharing with no restrictions outside what is stated in the Data Use Agreement.
- Cohort Principal Investigator (PI) is informed on how data is being used and the PI is responsible for informing participants on this usage when necessitated by local law/regulations/or IRB to do so
- Data has no intellectual property limitations, country policy, legal, or other restrictions on sharing and use of participant data, including but not limited to

- embargo period restrictions
- data use restrictions on combining data from multiple sources
- any restrictions that further restrict data use beyond the single AMP PD Data Use Agreement

BioFIND: All study protocols and recruitment strategies were approved by the institutional review boards for the University of Rochester Clinical Trials Coordination Center (CTCC) and individual sites. Tracking Parkinson’s: West of Scotland Research Ethics Service (WoSRES) Research Ethics Committee gave ethical approval for this study (ref 11/AL/0163). PPMI: The Research Subjects Review Board at the University of Rochester approved the PPMI study protocol. Oslo: the Regional Committee for Medical Research Ethics in South-East Norway gave ethical approval for this study. Participants’ information and genetic samples were obtained under appropriate written consent and with local institutional and ethical approvals. QPN: All patients recruited through the QPN have signed an informed consent form at enrollment, and the study protocol was approved by the institutional research ethics board.

## Author contributions

Concept and design: MMXT, LP

Acquisition, analysis or interpretation of data: MMXT, HI, SB-C, YS, KB, NMW, RNA, JM-G, GA, O-BT, PA, SE, PH, DKS, KK, CHW-G, DGG, J-CC, ZG-O, MT, LP

Drafting of the manuscript: MMXT, LP

Critical revision of the manuscript: MMXT, HI, SB-G, YS, MS, KB, NMW, RNA, JM-G, GA, O-BT, PA, SE, PH, DKS, KK, JH, CHW-G, DGG, J-CC, ZG-O, MT, LP

Statistical analysis and supervision: MMXT, SB-C, HI

