## Supplementary Materials for "Polygenic scores for disease risk are not associated with clinical outcomes in Parkinson’s disease"

### **Supplementary Methods**

#### AMP-PD cohorts

For six of the in-person cohorts, clinical and genetic data were accessed through version 2.5 of the Accelerating Medicines Partnership (AMP-PD) platform. These included the Harvard Biomarkers Study (HBS), Parkinson’s Progression Markers Initiative (PPMI), BioFIND, Parkinson’s Disease Biomarkers Program (PDBP), SURE-PD3, STEADY-PD3, and the LRRK2 Cohort Consortium (LCC). Only participants recruited with a diagnosis of PD at study entry were included. Participants who were initially recruited as individuals with PD but later rediagnosed with a non-PD condition were excluded (N = 36). We also excluded participants recruited in the genetically-enriched cohorts through the PPMI study, as these participants (both PD cases and controls) carry specific high-risk alleles such as *GBA1* p.N370S or *LRRK2* p.G2019S and, therefore this genetic enrichment could bias analyses^1^.

BioFIND^2^: This study is a cross-sectional observational study investigating biomarkers in PD, which took place at multiple sites across the US. PD participants met the modified UK PD Society Brain Bank (UKPDBB) clinical diagnostic criteria, requiring all three classic motor signs of parkinsonism: bradykinesia, rigidity, and resting tremor. Participants were required to have progressive PD of 5 to 18 years of duration from the onset of motor symptoms. The age of onset was required to be between 50 and 75. Individuals were excluded if they had any other serious neurological or psychiatric disorder, a history of a major medical condition (e.g. cancer or liver disease), early onset autonomic symptoms, prior brain surgery (including Deep Brain Stimulation), or if they were taking an investigational drug.

HBS (<https://www.bwhparkinsoncenter.org/biobank/>^3^): The Harvard Biomarkers Study (HBS) is a biobank and longitudinal case-control study of individuals with PD and other neurodegenerative disorders. PD participants were required to be 21 years or older, have an MMSE > 21 or provide joint consent with a primary caregiver if they had known dementia or MMSE below 21, and a diagnosis of PD.

LCC^4^: The LRRK2 Cohort Consortium (LCC) is a multi-site study of PD and parkinsonism and their family members. The study recruited idiopathic PD patients, PD participants carrying a mutation in the *LRRK2* gene, unaffected subjects carrying a *LRRK2* mutation, and healthy control subjects. We only included idiopathic PD participants in our analysis and not participants in other study arms. Idiopathic PD participants were eligible if they were of Ashkenazi Jewish descent and had PD or parkinsonism, and were over 18 years old.

PDBP (<https://pdbp.ninds.nih.gov/>^5^): The Parkinson's Disease Biomarkers Program (PDBP) aims to facilitate biomarker research and is a longitudinal observational study. PD participants were required to be clinically diagnosed with PD, aged over 21 years at screening, and able to cooperate with consent procedures and study activities. Participants were excluded if they were treated with anticoagulants, had a history of neuroleptic use or exposure, had a history of schizophrenia, used investigational drugs or devices within 60 days prior to their baseline visit, or were otherwise unable to participate in biological sample collection, routine lumbar puncture or clinical assessments.

PPMI (<https://www.ppmi-info.org/>;^6^): The Parkinson’s Progression Markers Initiative (PPMI) is a longitudinal, observational and multicentre study. For PD subjects, participants were required to have at least two of the following: resting tremor, bradykinesia, rigidity (must have either resting tremor or bradykinesia), or either asymmetric resting tremor or asymmetric bradykinesia. Participants had to have a diagnosis of PD for 2 years or less at screening, aged 30 years or older at time of PD diagnosis, Hoehn and Yahr stage I or II at baseline, confirmation of dopamine transporter deficit from dopamine transporter SPECT scan, and not expected to require PD medication within at least 6 months from baseline. PD participants were ineligible if they were taking a PD medication, had taken levodopa, dopamine agonists, MAO-B inhibitors or amantadine within 60 days of Baseline, or had taken levodopa or dopamine agonists prior to Baseline for more than a total of 60 days. Participants were also excluded if they received any drugs that might interfere with DaTSCAN imaging, treated with anticoagulants, had a condition that interfered with safe performance of routine lumbar puncture, or used an investigational drug or device within 60 days prior to baseline. There are also other study arms within PPMI (healthy controls, prodromal subjects and genetic cohorts) however we only analysed the PD subjects.

SURE-PD3^7^: This is a randomised double-blind trial of urate-elevating inosine treatment to slow clinical decline in early PD. Participants were required to fill diagnostic criteria for idiopathic PD with at least two of the cardinal signs of PD (resting tremor, bradykinesia, rigidity), not have current or imminent (within 90 days of enrollment) PD disability requiring dopaminergic therapy, have a modified Hoehn and Yahr scale stage 1 to 2.5 inclusive, aged 30 or older at the time of PD diagnosis, diagnosis of PD within 3 years prior to the first screening visit, non-fasting serum urate ≤ 5.7 mg/dL at first screening visit. Female participants had to be either surgically sterile, postmenopausal or be using a reliable form of contraception and negative pregnancy test at screening. Participants were also required to have a MMSE score ≥ 25.

STEADY-PD3 was not included in analyses as age at diagnosis was missing for all participants in the AMP-PD Terra portal. The LBD was also not included as clinical data was not available. Further information about AMP-PD and the cohorts can be found at <https://amp-pd.org/>.

#### Oslo

The Oslo PD cohort is a registered biobank for patients with parkinsonism at Oslo University Hospital. PD participants in the Oslo study were recruited from a tertiary-care unit for movement disorders at Oslo University Hospital^8^. Participants were required to have a clinical diagnosis of PD to be eligible. Notably, a large proportion of referrals to the unit were for evaluation for advanced treatment options, such as Deep Brain Stimulation (DBS). Thus the patient group in the study may have an earlier age at onset, severe motor fluctuations, good levodopa response and better cognitive function than other PD cohorts^8^. The study was approved by the Regional Committee for Medical Research Ethics in South-East Norway.

#### Quebec Parkinson’s Network

The Quebec Parkinson’s Network (QPN) is an open-access patient registry and biosample repository. It is an observational, multicenter longitudinal study^9^. Participants were recruited at tertiary and academic centres in the province of Quebec. PD participants were eligible if they were diagnosed by a movement disorder specialist in Quebec according to the MDS criteria or UK Brain Bank Criteria. All participants provided informed consent at study entry and the study was reviewed and approved by the institutional research ethics board.

#### Tracking Parkinson’s

Tracking Parkinson’s is a prospective, longitudinal, multi-centre observational study recruiting patients from secondary care health centres across the UK^10^. PD participants in the longitudinal study arm were required to be diagnosed with PD within 3.5 years of study enrollment (recent onset PD cases). PD participants were recruited if they were aged 18 to 90 years and had a clinical diagnosis of PD according to the Queen Square Brain Bank criteria and supported by neuroimaging if the clinical diagnosis was uncertain. Patients could be drug naïve or treated. Participants were excluded if they had severe comorbid illness that restricted participation in clinic visits, or other degenerative forms of parkinsonism.

#### NIH cohorts

DATATOP: DATATOP (Deprenyl and tocopherol antioxidative therapy of parkinsonism) is a randomized clinical trial of deprenyl 10g/d and/or tocopherol 2000 IU/d in early PD patients^11^. The study took place at 28 US and Canadian sites and enrolled 800 untreated PD patients. PD patients were eligible if they were assessed to have early PD (stage 1 and 2), disease duration ≤ 5 years, aged between 30 and 79 years, and were not receiving or requiring any anti-PD medications. Participants were systematically evaluated at regular intervals over 2 years.

DIGPD: Drug Interaction with Genes in Parkinson's Disease (DIGPD) is a multicentre longitudinal observational cohort of PD patients followed for up to 5 years^12^. PD participants were required to have disease duration ≤ 5 years at baseline and meet the UK Parkinson’s Disease Society Brain Bank criteria. Participants were recruited from French university hospitals and general hospitals between 2009 and 2013. At the time that clinical and genetic data was shared with the National Institutes of Health, the cohort was divided into two according to the genotyping array that samples were genotyped with: either the NeuroX (a targeted chip for neurodegenerative disease) or the Illumina Mukti-Ethnic Genotyping Array.

PARKFIT: The ParkFit study is a multicenter, randomised controlled trial of physical therapy in PD patients in the Netherlands^13^. PD participants were eligible if they had PD according to the UK Brain Bank Criteria, aged between 40 and 75 years, had a sedentary lifestyle defined as < 3 times a week vigorous-intensity physical activity for < 60 minutes; or < 3 times a week moderate-intensity physical activity for < 150 minutes), and had a Hoehn and Yahr stage of 3 or less. Participants were excluded if the diagnosis was unclear (no gratifying and sustained response to dopaminergic therapy), MMSE < 24, unable to complete Dutch questionnaires, severe comorbidity, daily institutionalised care, or deep brain surgery. 586 participants with PD were randomised to either a multifaceted program of behavioural change to promote physical activity, or physical therapy interventions.

PARKWEST: The Norwegian ParkWest study is a prospective longitudinal study of incident PD in four Norwegian counties in Western and Southern Norway^14^. The study sought to recruit all residents with incident PD in the counties of Sogn and Fjordane, Hordaland, Rogaland, and Aust-Agder between November 2004 and August 2006. PD participants were required to have at least two of the four cardinal motor signs, typical disease history with evidence of progressive parkinsonism, no dementia at parkinsonism onset, and no severe atypical signs. Dopamine transporter imaging was also used to help with diagnosis. 212 incident PD participants were recruited and followed up longitudinally.

PICNICS: The Parkinsonism: Incidence and Cognitive and Non-motor heterogeneity In CambridgeShire (PICNICS) is a prospective longitudinal study of patients with newly-diagnosed PD recruited in Cambridgeshire, UK, between April 2008 and 2013^15^. Participants were recruited through normal routes of UK/National Health Service (NHS) healthcare referral, where general practitioners and hospital specialists in the county were asked to refer consenting cases of new or suspected PD or parkinsonism to the study investigators. Idiopathic PD was diagnosed with the Queen Square Brain Bank criteria. 280 participants with PD were recruited and followed up longitudinally every 18 months.

PRECEPT^16^: The Parkinson Research Examination of CEP-1347 Trial (PRECEPT) is a multicentre, randomised, placebo-controlled trial of CEP-1347 as a potential disease modifying treatment in PD. CEP-1347 acts to inhibit mixed lineage kinases that survive apoptotic pathways thought to be important for PD pathogenesis^16^. Patients with early PD who were untreated and had not yet developed disability requiring dopaminergic treatment. The primary endpoint of the study was the time to development of disability requiring dopaminergic therapy. PD patients were required to be older than 30 years, have two of the cardinal signs of PD, modified Hoehn and Yahr stage ≤ 2.5, and no current or imminent disability requiring dopaminergic therapy. Patients were excluded if they had atypical or drug-induced parkinsonism, a diagnosis of PD ≥ 5 years duration, a rest tremor score ≥ 3 in any limb, had used antiparkinsonian therapy within the past 6 months, history of malignant melanoma, history of cancer, history of seizures, MMSE ≤ 26, Beck depression score ≥ 15, creatinine clearance ≤ 50 mL/min, or other trial participation or medication use which may interact with CEP-1347. 806 participants with early PD were enrolled at 65 sites across the United States and Canada. Though the trial terminated early due to futility, a proportion of patients were continuously followed-up in an observational study called PostCEPT^17^.

#### Acknowledgements

We thank all of the study participants and their families and the investigators and members of the following studies: Parkinson Study Group: Deprenyl and Tocopherol Antioxidative Therapy of Parkinsonism (DATATOP); Drug Interaction with Genes in Parkinson’s Disease (DIGPD); Harvard Biomarkers Study (HBS); NET-PD_LS1, National Institutes of Health Exploratory Trials in Parkinson’sDisease Large Simple Study 1; Oslo PD study; ParkFit Study; The Norwegian ParkWest Study (PARKWEST); Parkinson’s Disease Biomarker Program (PDBP); Parkinsonism Incidence and Cognitive and Non-motor heterogeneity In Cambridgeshire (PICNICS); Parkinson’s Progression Markers Initiative (PPMI); Parkinson Study Group: Parkinson Research Examination of CEP-1347 Trial (PreCEPT) and its following study (PostCEPT); Profiling Parkinson’s Disease Study (PROPARK); and Morris K. Udall Centers for Parkinson’s Research (Udall). We also thank the several grants and financial supporters of the previous studies. DATATOP was supported by a Public Health Service grant (NS24778) from the National Institute of Neurological Disorders and Stroke (NINDS) and by grants from the General Clinical Research Centers Program of the National Institutes of Health at Columbia University (RR00645), the University of Virginia (RR00847), the University of Pennsylvania (RR00040), the University of Iowa (RR00059), Ohio State University (RR00034), Massachusetts General Hospital (RR01066), the University of Rochester (RR00044), Brown University (RR02038), Oregon Health Sciences University (RR00334), Baylor College of Medicine (RR00350), the University of California (RR00827), Johns Hopkins University (RR00035), the University of Michigan (RR00042), and Washington University (RR00036), the Parkinson’s Disease Foundation at Columbia-Presbyterian Medical Center, the National Parkinson Foundation, the Parkinson Foundation of Canada, the United Parkinson Foundation, Chicago, the American Parkinson’s Disease Association, New York, and the University of Rochester. DIGPD is supported by Assistance Publi- que Hôpitaux de Paris and funded by a grant from the French Ministry of Health (PHRC 2008, AOM08010) and a grant from the Agence Nationale pour la Sécurité des Médicaments (ANSM 2013). HBS is supported by the Harvard NeuroDiscovery Center, Michael J. Fox Foundation, NINDS U01NS082157, U01NS100603, and the Massachusetts Alzheimer’ Disease Research Center NIA P50AG005134; National Institutes of Health Exploratory Trials in Parkinson’s Disease Large Simple Study 1 was supported by NINDS Grant U01NS043128. OSLO is supported by the Research Council of Norway and South-Eastern Norway Regional Health Authority. ParkFit is supported by ZonMw (the Netherlands Organization for Health Research and Development [75020012]) and the Michael J. Fox Foundation for Parkinson’s Research, VGZ (health insurance company), GlaxoSmithKline, and the National Parkinson Foundation. ParkWest is supported by the Research Council of Norway, the Western Norway Regional Health Authority, Stavanger University Hospital Research Funds, and the Norwegian Parkinson’s Disease Association. PDBP is a consortium with NINDS initiative. PICNICS has received funding from the Cure Parkinson’s Trust and the Van Geest Foundation and is supported by the National Institute of Health Research Cambridge Biomedical Research Centre. PPMI is supported by the Michael J. Fox Foundation for Parkinson’s Research. PreCEPT and PostCEPT were funded by NINDS 5U01NS050095-05 and the Department of Defense Neurotoxin Exposure Treatment Parkinson’s Research Program, Grant W23RRYX7022N606, the Michael J. Fox Foundation for Parkinson’s Research, Parkinson’sDis- ease Foundation, Lundbeck Pharmaceuticals, Cephalon Inc, Lundbeck Inc, John Blume Foundation, Smart Family Foundation, RJG Foundation, Kinetics Foundation, National Parkinson Foundation, Amarin Neuroscience LTD, CHDI Foundation Inc, National Institutes of Health (NHGRI, NINDS), and the Columbia Parkinson’s Disease Research Center. PROPARK is funded by the Alkemade-Keuls Foundation, Stichting Parkinson Fonds, Parkinson Vereniging, and The Netherlands Organisation for Health Research and Development. Udall is supported by NINDS.
